# The Rapid Deployment of a 3D Printed “Latticed” Nasopharyngeal Swab for COVID-19 Testing Made Using Digital Light Synthesis

**DOI:** 10.1101/2020.05.25.20112201

**Authors:** Ian Bennett, Philip L. Bulterys, Melody Chang, Joseph M. DeSimone, Jennifer Fralick, Marie Herring, Hardik Kabaria, Christina S. Kong, Blane Larson, Owen Lu, Arielle Maxner, Elle Meyer, Shawn Patterson, Steven Pollack, Jason Rolland, Steven Schmidt, Sridhar Seshadri, Keshav Swarup, Chelsey Thomas, Ryan Van Wert

## Abstract

The novel coronavirus disease (COVID-19) has caused a pandemic that has disrupted supply chains globally. This black swan event is challenging industries from all sectors of the economy including those industries directly needed to produce items that safeguard us from the disease itself, especially personal protection equipment (N95 masks, face shields) and much needed consumables associated with testing and vaccine delivery (swabs, vials and viral transfer medium). Digital manufacturing, especially 3D printing, has been promulgated as an important approach for the rapid development of new products as well as a replacement manufacturing technique for many traditional manufacturing methods, including injection molding, when supply chains are disrupted. Herein we report the use of Digital Light Synthesis (DLS) for the design and large-scale deployment of nasopharyngeal (NP) swabs for testing of coronavirus SARS-CoV-2 infections in humans. NP swabs have been one of society’s essential products hardest hit by the supply chain disruptions caused by COVID-19. A latticed tip NP swab was designed and fabricated by DLS from a liquid resin previously developed and approved for use to make dental night guard devices. These latticed NP swabs demonstrated non-inferiority in a human clinical study of patients suspected of being infected with SARS-CoV-2.

## INTRODUCTION

The novel coronavirus disease 2019 (COVID-19) is caused by the newly emerged, highly contagious coronavirus SARS-CoV-2 which invades the respiratory tract and the lungs, as well as other organ systems. Rapid detection of viral infection is essential for accurate tracking and mitigation of epidemic spread (1). A major obstacle hindering the large-scale rollout of COVID-19 tests is the lack of nasopharyngeal (NP) swabs used for collecting viral samples. One of the largest manufacturers of NP swabs is located in Lombardy, Italy, which is one of the areas in Europe hardest-hit by COVID-19 (2). In response to this national shortage, we set out to design an NP swab suitable for manufacture using Digital Light Synthesis™ (DLS) (3), and to clinically evaluate its performance. This multidisciplinary initiative to address the national shortage of NP swabs began in partnership with an *ad hoc* consortium out of Beth Israel Deaconess Medical Center (BIDMC) and Harvard University (4).

## RESULTS

### Design / Problem Statement

The NP swab has three important functions: 1) it must easily and comfortably pass through the highly curved nasal cavity; 2) it must collect a sufficient mucosal sample to test for viral RNA; and 3) it must release the biological sample in a manner that will not interfere with the reverse transcription polymerase chain reaction (RT-PCR) test.

### Latticed NP Swab Design

Considering these factors, we split the design of the NP swab into two important parts: 1) the bulb tip at the distal end and 2) the handle. We began by designing the overall length and handle dimensions to closely mimic the standard flocked NP swabs. This included a breakpoint on the handle located at 70 mm from the distal end that allows clinicians to break off the swab bulb and deposit the collected sample into standard sized sample tubes. We introduced a concept of the latticed bulb at the distal end to replace the flocked tip in a traditional NP swab. The reason behind using a lattice structure as the bulb tip was to create a flexible tip (for comfort and nasal cavity passage) with a similar exterior dimension (diameter of 2.6 mm) while at the same time having open cavities to facilitate the collection and transfer of biological sample.

To design a latticed swab to replace the traditional flocked swab, we focused on optimizing the two important parameters of the design: 1) the shape of the bulb tip, and 2) the structure of the lattice that fills the shape of the bulb. The design iterations were guided by manufacturing process requirements and feedback from clinicians at Stanford University. Our goal was to optimize for sufficient sample collection as well as patient comfort. These design iterations converged to a 35 mm long conical shape bulb tip of the swab and lattice parameters that led to an as open as possible, yet conformal, lattice structure of the bulb. The latticed bulb is designed to capture and retain a large volume of sample through a combination of surface energy and geometry. This design has been developed in part using the lattice design software (10) developed at Carbon, Inc.

Based on the first clinical trial at BIDMC (4) and the patient feedback pertaining to comfort relative to a conventional flocked swab, we introduced a Generation 2 latticed NP swab design with two main changes: a fully tapered, helical core inside the bulb tip and a more open unit cell design with a smoother lattice top to increase patient comfort during sampling. These modifications improved durability and flexibility in the bulb, while maintaining sample collection volume.

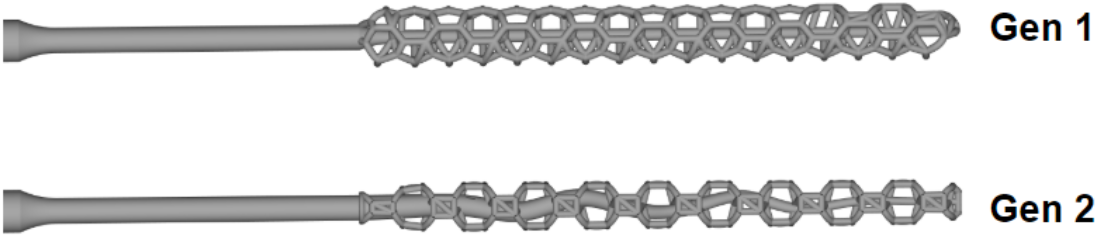

The tapered, helical core pre-bends the bulb tip region, allowing it to flex when minimal force is applied, and to more easily conform to the contours of the nasal cavity. The more open unit cell also improves flexibility by removing many of the axially oriented struts. Additionally, the larger unit cell allows biological sample to more easily enter the lattice cage where it remains captured for collection.

### Material Selection

NP swabs require a combination of flexibility and strength for safe and effective sample collection. In general, the swab must be able to bend backward on itself without breaking, while still having sufficient rigidity to reach the nasopharynx, pushing through the drag of the nasal passageway. The material that makes up the swab bulb must be sufficiently porous and soft to be comfortable, without any weakness that could result in a fracture during sample collection. In order to achieve these results, we postulated that the material for the latticed NP swab needed to have a relatively high elongation at break (above 50 %) and a Young’s modulus above 500 MPa. For a medical device in contact with mucosa, the requirements are that the final finished device pass cytotoxicity, irritation (ISO 10993-5 and −10) and sensitization biocompatibility testing requirements after sterilization. Given the urgent timeframes associated with responding to the COVID-19 pandemic we felt that it would be ideal to utilize an existing resin suitable for printing via DLS that was already approved for use in the dental marketplace and had the requisite mechanical and biological performance properties. We also evaluated which resins of those previously validated for DLS were compatible with autoclave sterilization as well as other potential sterilization methods. The KeySplint Soft® Clear material from Keystone® Industries met these criteria.

### Clinical Assessment of Suspected and Known SARS-CoV-2 Infected Patients

At Stanford University, the latticed NP swabs were clinically validated on suspected SARS-CoV-2 infected patients through an IRB approved study. The performance of the latticed NP swab was compared to the Copan model 501CS01 (“control”) NP swab. Participants scheduled for standard clinical SARS-CoV-2 RT-PCR testing with a control swab were consented for a second sample using the latticed NP swab. Sample collection was performed by staff who underwent formal training in obtaining NP swab samples. Data from testing of 176 study participants at Stanford Hospital using the Generation 1 latticed NP swabs are shown in Figure 1. The combination of the absence any true positive (TP) or false positive (FP) calls precludes Kappa coefficient calculation for these data. Data from testing of 201 study participants at Stanford Hospital using the Generation 2 latticed NP swab are shown in Figure 1.

**Figure 1:**
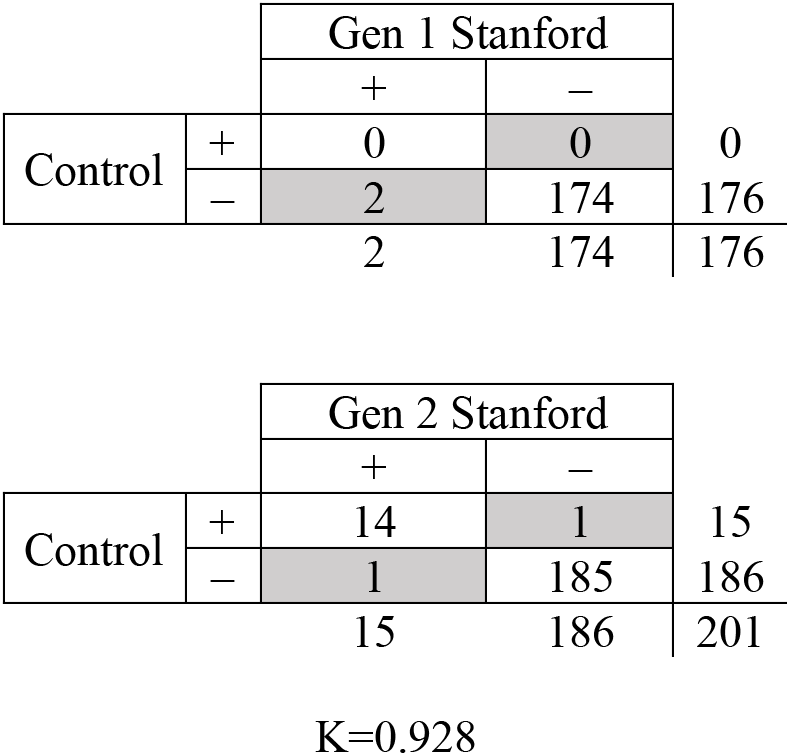
Concordance results shown as 2×2 tables giving counts for Generation 1 and 2 latticed NP swab vs. the control swab. Discordant results in gray; totals for each swab below and to the right of each box; K=Cohen’s kappa.

Kappa coefficients suggested a high degree of agreement (7,8) between the latticed NP swabs and the traditional flocked swabs from COPAN. It should be noted that for the reading ranked as a False Positive, this assignment was because the flocked swab sample was read as negative. However, it was determined that the patient from whom this sample was obtained was symptomatically COVID-19 positive, and had tested COVID-19 positive twice in the weeks prior using traditional flocked swabs. We hypothesize that this false negative for the flocked swab sample was due to a low viral burden for this patient (9) and could be attributed to lower viral RNA recovery for the flocked swab relative to the latticed NP swab.

### Conclusions

The DLS technology enabled the rapid design, scale up and clinical deployment of an entirely new class of a much-needed medical device—the NP testing swab—during the COVID-19 pandemic. DLS allowed multiple iterations of the latticed NP swab, including clinical evaluation in humans, in a very short period of time. The latticed NP swabs were shown to be equivalent or better compared to the flocked swabs (from COPAN) based on the clinical study with almost 400 patients. A significantly different physical structure— a latticed bulb tip versus a flocked swab—was found to be suitable to collect a biological sample in the nasopharynx. The KeySplint Soft® Clear material from Keystone® Industries was found to be suitable for use as a latticed NP swab with demonstrated compatibility with RT-PCR analytical methods to detect the coronavirus SARS-CoV-2. Using Carbon’s additive manufacturing method, DLS, along with the right choice of material, it was possible to design, clinically validate, and launch a Class I, 510(k) Exempt *in vitro* diagnostic medical device within just 20 days.

Additively manufacturing NP swabs emerged to address the critical shortages brought about by the COVID-19 pandemic. Understanding the strengths and shortcomings of these digitally designed latticed test swabs will require further study under more controlled, multi-center studies. The lower apparent rate of false negatives using the latticed NP swabs is encouraging, but will need to be examined in a larger clinical study. Recent work has shown (9) that at different phases of the disease, the nasopharyngeal site may not be optimal for identifying COVID-19 positive patients. Designing swabs optimized for mid-turbinate or nares collection and comparing the relative accuracy of these sample sites is a potential area of study. Test validation in these less “invasive” anatomical sites may allow for a better patient experience and the potential for self-administered sampling, helping to lower the risk to healthcare workers and patients.

## MATERIALS AND METHODS

The latticed NP swabs were 3D printed using DLS technology on M2 printers (192 mm × 120 mm XY build area with a 75 micron pixel size) using the KeySplint Soft® Clear resin. The latticed NP swabs were printed at a packing density spaced approximately 6 mm apart on center which enabled 448 swabs to be printed in 2 hours 30 minutes. Manufacturing process instructions were developed in concordance to the *Indications for Use* associated with Keystone® Industries’ FDA 510(k) filing K183598 for the KeySplint Soft® Clear resin. These were largely followed in order to leverage the corresponding biocompatibility data. To improve yield and throughput, a sacrificial printed feature was added to the part file to minimize part handling. This feature on the parts also encompassed a unique identifier for traceability throughout numerous manufacturing and shipping processes. After printing, the latticed NP swabs were washed with >97% isopropyl alcohol for 5 minutes. Next, they were left to air dry for 30 minutes. Once dry, the swabs were UV-cured for 20 minutes in a Dreve PCU 90 post-cure unit under a nitrogen inert environment using a custom fixture for stacking within the UV chamber. The swabs were then inspected according to quality plan requirements before being individually packaged in a single-use autoclavable package that was shipped to the clinical sites.

The latticed NP swabs were tested after sterilization using a panel of laboratory and clinical tests to determine performance according to identified metrics including mechanical properties (tensile strength, torque, simulated-use), collection efficiency (water uptake), PCR compatibility and clinical performance.

### Mechanical Properties

The latticed NP swabs were tested to failure using an MTS® Tester (MTS Exceed E43) to determine the force required to separate the latticed bulb tip from the handle. This was to ensure that the latticed NP swab could withstand forces beyond what it might experience *in situ*. From a practical viewpoint, there is very little evidence that NP swabs will get caught on a protuberance in the nasal cavity. A 4.5 N load was selected as a minimum requirement as it would allow a significant safety factor over what would be experienced by the swab through interaction with the mucus membrane and a force that could be reasonably expected during collection by a healthcare worker. The latticed NP swabs were twisted 90° when loaded into the gripper of the MTS tester and pulled to failure. The purpose of the 90° twist was to expose any flaws or weak points in the neck. Table 1 shows that both Generation 1 and Generation 2 swabs withstood over 25 N before breakage, over the ISO 10993-5 and −10 subparts which correspond to 5 times the maximum expected load. To understand what a reasonable force the NP swab could expected to encounter when twised during collection and to see if that force would cause the latticed bulb tip to separate from the handle, the NP swabs were analyzed by a torque tester. A gloved hand was used to apply rotational force to the handle and the maximum torque was measured. It was not possible to apply more than 2.7 Nm of torque to the NP swab. This force was not enough torque to separate the latticed bulb tip from the handle and all samples survived without failure.

**Table 1:** Results from mechanical testing and water uptake on the latticed NP swabs.

|  | Tensile Testing | Simulated Use Testing | Water Uptake |
| --- | --- | --- | --- |
| Gen 1 | 33.45 ± 6.81 N | Pass (n=22) | 89.27 ± 15.56 µL |
| Gen 2 | 26.99 ± 4.93 N | Pass (n=22) | 61.28 ± 3.05 µL |

To further understand the performance of the latticed NP swab, a simulated use test was developed that used a section of clear, flexible ¼” inner diameter tubing bent with a radius of 28 mm in a fixture. The testing was done with no lubrication as a worst case (as compared to using mineral oil). To test the latticed NP swab, the distal end was inserted into the tubing until the bulb reached the bend of the tubing, noted by a landmark on the fixture. While in this position, the swab was turned 5 times clockwise, then 5 times counterclockwise, then examined for breakages. Swabs that did not experience multiple strut failures were deemed as passing. Results from this test can be found in Table 1. Separately, large numbers of latticed NP swabs (>600 of each) were tested in simulated use for process validation and were observed for the first single strut breakage. Generation 1 samples were able to get to 10 turns, on average, while many (N=20) Generation 2 swabs maxed out at 21 turns (upper limit of testing) with no breakages. Results are shown in Figure 2 indicating that both designs clearly withstand worse case testing.

**Figure 2:**
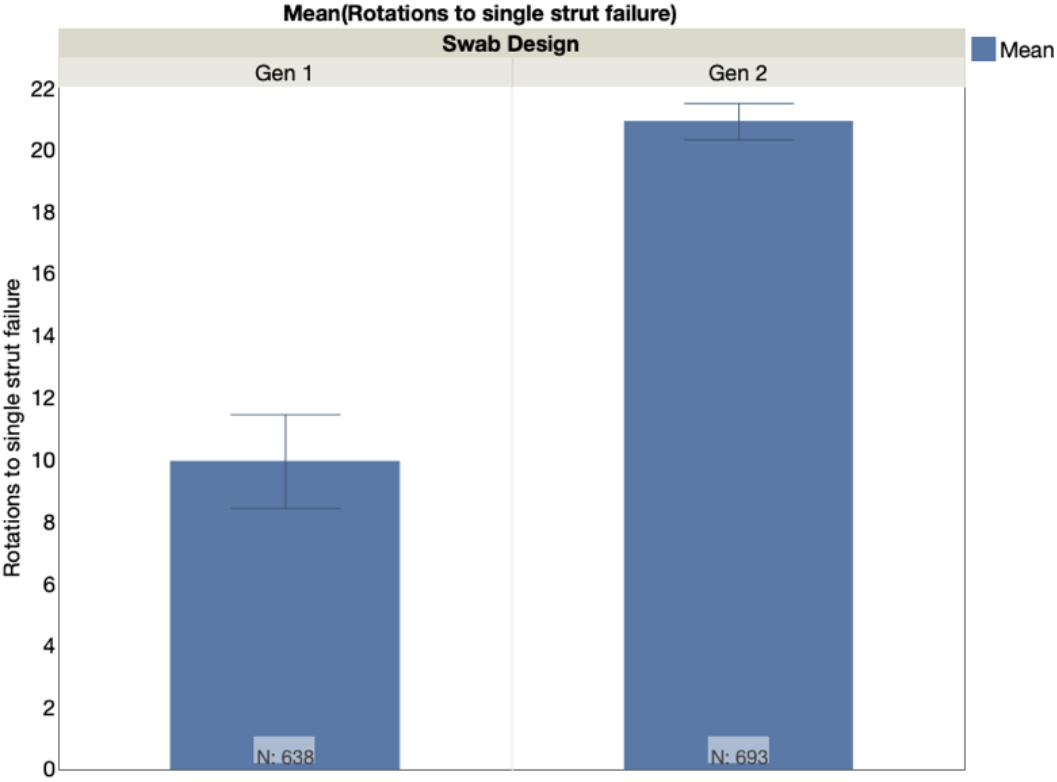
In process validation, a large number (>600) of samples were tested in simulated use models. The samples were monitored carefully for the first sign of a single strut failure. Compared to Generation 1 swabs, Generation 2 swabs were able to withstand almost double the turns until the first strut failure.

The latticed NP swabs were tested for bending force by using a 3-point bend fixture. The latticed bulb tip was placed across two fixed posts and the force was tested to displace the middle of the latticed bulb tip by 5 mm. Figure 3 shows the curve of comparative load at equivalent extensions for Generation 1 and Generation 2 latticed NP swabs. Because of the improved flexibility of the Generation 2 latticed NP swabs, much less force was required to deflect the latticed swab bulb tip, presumably facilitating navigation through non-linear nasal passageways. The difference in bending modes between the swabs is shown in Figure 4. When a load is applied to the bulb tip, the Generation 1 latticed NP swab bent mostly at the neck, below the swab, while the Generation 2 latticed NP swab bent mostly in the swab bulb tip.

**Figure 3:**
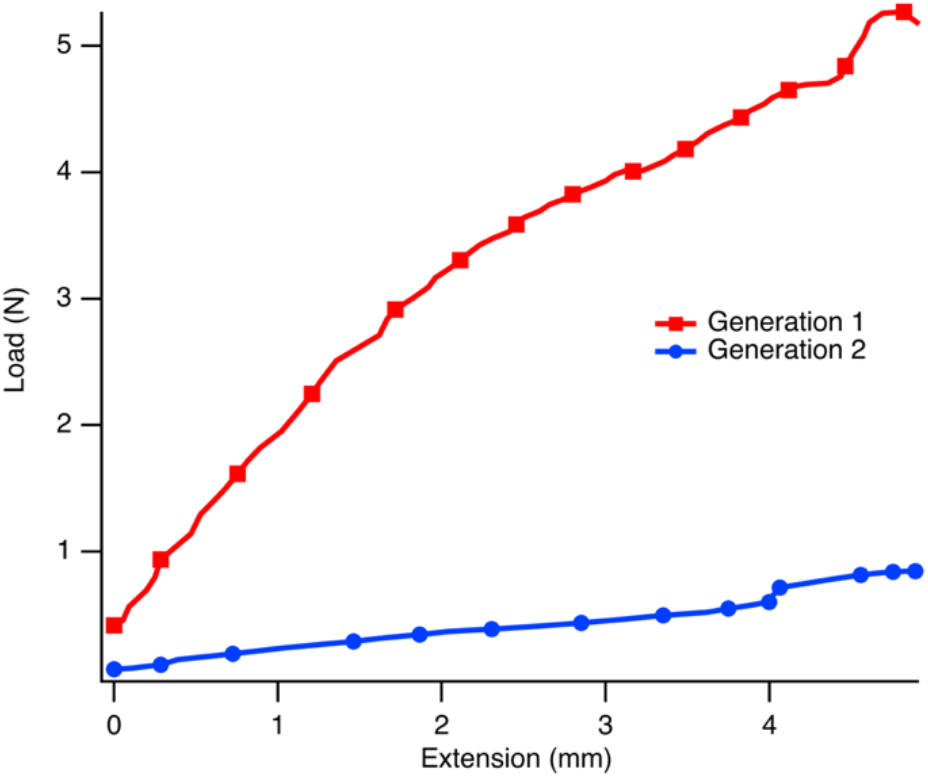
Comparison of Generation 1 and Generation 2 latticed NP swabs in the three-point bend force measurement test. Generation 2 requires less force to deflect, which allows it to navigate the nasal passageway with less force.

**Figure 4:**
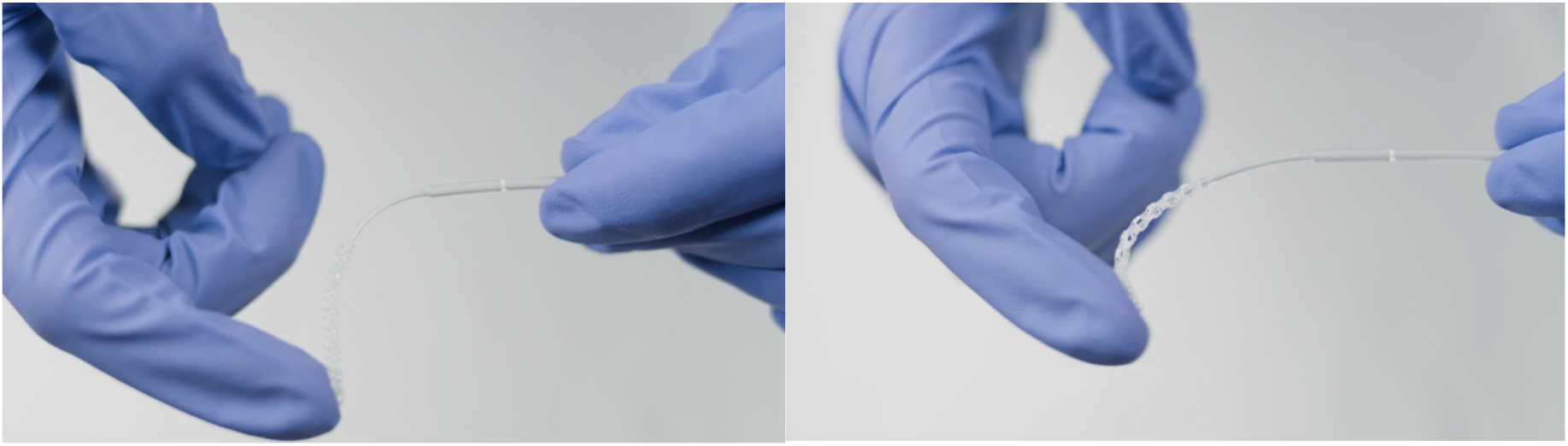
Comparison of bending modes for Generation 1 (Left) and Generation 2 (Right) latticed NP swabs. When force is applied to the tip of the swab, Generation 1 reacts by bending the most in the neck, below the bulb. In Generation 2, the bending is concentrated in the swab bulb.

### Collection Efficiency

The main goal of a NP swab is to collect enough secretions to accurately detect viral particles by RT-PCR. As a proxy for sample collection, we tested water uptake of the latticed NP swabs. Water (1 mL) was added to an analytical scale and the swab’s surface was then completely exposed (rolled) in the water. The water uptake was determined gravimetrically. Both Generation 1 and Generation 2 swabs were able to uptake a minimum of 50 μL of water, far exceeding amounts of water uptake reported (16 μL) for traditional NP flocked swabs (5).

### PCR Compatibility

For this application, in addition to suitable mechanical and collection efficiency properties, it is of utmost importance that the latticed NP swabs do not interfere with the PCR assay. It is well known that swabs made from natural fiber sources (cotton-tipped or alginate treated) can interfere with the assay either by strongly binding the RNA/DNA analyte or by inhibiting the enzymatic reactions in the test reagents (6). Given that the latticed NP swabs were being made using a material of unknown compatibility with PCR testing, they were evaluated with several different analytical systems including the Qiagen QIAsymphony SP and Abbott’s M2000 Aldatu test. No inhibition of PCR due to the material of the swab or any other factor was detected.

The latticed NP swabs used for clinical assessment were manufactured by Resolution Medical a U.S. based, FDA registered, *in vitro* diagnostic and medical device manufacturer who is in compliance with all applicable regulations, including the Quality System Regulation 21 CFR Part 820 and ISO 13485:2016. Resolution Medical is a licensed customer of Carbon and has multiple Carbon printers. The latticed NP swab is a Class I 510(k) Exempt, *in vitro* diagnostic medical device registered with the FDA (#10050670). The latticed NP swabs were individually packaged and autoclaved for sterilization according to manufacturer protocols prior to use (Sterilizer Type: Prevacuum; Preconditioning Pulses: 4; Temperature: 132-degrees Celsius; Full Cycle Time: 4 minutes; Dry Time: 30 minutes). Swabbing was performed per standard protocol. Participants were first swabbed with the control swab, then the latticed NP in the opposite nostril. Choice of naris for each swab was left to staff member and participant. Control and test NP swabs were placed in separate vials of buffered saline and tested with a lab developed SARS-CoV-2 RT-PCR assay that received FDA emergency use authorization (EUA200036). RT-PCR results are reported as positive, negative, or indeterminate. For purposes of calculating Cohen’s Kappa (7,8), the flocked swabs were assumed to be the gold standard (although it is reported that the flocked swabs demonstrate < 100% accuracy (4)). It must be noted that it is less desirable for the clinical sites to sterilize the swabs locally prior to use, however there are numerous quality assurance tests associated with shipping pre-sterilized products whose timelines were not conducive to a rapid response in the context of the COVID-19 pandemic. A version of the latticed NP swab sterilized using ethylene oxide is currently under development and should be deployed by June 2020.

## Data Availability

All of the data is included directly in the manuscript.

## Acknowledgements

We thank R. Fulop, K. Parker and R. Arnaout at Desktop Metal, Harvard University and Beth Israel Deaconess Medical Center, respectively, for bringing to our attention the awareness that there was a national shortage of NP testing swabs and for standing up an *ad hoc* consortium of 3D printing experts to address the COVID-19 diagnostic testing shortages (https://github.com/rarnaout/Covidswab). We would like to sincerely thank all of the patients who participated in this study. In addition, we acknowledge the efforts of the Stanford care providers, in particular Julia A. Bancroft, MSN, FNP-C, who made this study possible.

## Funding

We thank Stanford Health Care for providing the resources and funding for the study.

## Author Contributions

J.M.D, H.K., C.K., E.M., S.P., S.P., J.R., S.S. (Schmidt), S.S. (Seshadri), and R.V.W. performed the conceptualization, investigation, data analysis and writing. I.B., P.B., M.H., B.L., O.L., A.M., G.R., and K.S. assisted with the conceptualization, laboratory investigations, data collection, analysis and writing. M.C., J.F., and C.T. assisted with the clinical investigations. All authors approved this manuscript.

## Competing interests

All authors affiliated with Carbon are compensated by and have an equity stake in Carbon, Inc. All authors affiliated with Resolution Medical are compensated by and have an equity stake in Resolution Medical, LLC.

## Notes

### Clinical Trial

This study was reviewed and approved by the Stanford Institutional Review Board under protocol #55823. This was a non-interventional study with a Class 1 exempt medical device that did not assess participant health outcomes.

### Funding Statement

No external funding was used for this study.

